# Updated U.S. census benchmark sleep dataset v1.1

**DOI:** 10.64898/2025.12.27.25343087

**Authors:** Adam M. Jones, Bhavin R. Sheth

## Abstract

We previously documented and released a benchmark dataset for machine learning research on sleep stage classification [1]. Subsequently, it was pointed out in a preprint [2] that some recordings in the National Sleep Research Resource [3] include only binary wake-sleep annotations, instead of full sleep stage scoring using the Rechtschaffen and Kales (R&K) [4] or American Academy of Sleep Medicine (AASM) [5] standards. Because wake-sleep labels are an ontological mismatch and not just label noise, they do not belong in a dataset designed for full sleep stage classification. Therefore, we have updated our benchmark dataset (henceforth known as benchmark v1.0) to replace 16 recordings with suitable recordings from age- and sex-matched subjects, while all other dataset selection criteria and distributions have been preserved. Additionally, the total number of recordings and the composition of the training, validation, and testing sets remain unchanged. While this update is a minor revision, we want to distinguish its use from v1.0, and therefore have titled this update as benchmark v1.1. The file listings are provided on the GitHub repository (https://github.com/adammj/ecg-sleep-staging/).

## 1. Introduction

At the time of our original paper [1], machine learning research on sleep stage classification lacked a standardized benchmark dataset, making comparisons between human scorers and machine learning models difficult and often misleading. Studies typically used different recordings— even when drawing from the same public sources—with unreported inclusion criteria and scorer-specific practices that introduced substantial variability in reported agreement metrics. To address this, we introduced a large, diverse benchmark dataset curated from multiple source studies using a fully documented, automated selection pipeline to minimize sampling bias. We also released the exact recording lists to enable reproducible training, evaluation, and testing in future work.

The purpose of this update is to correct an oversight. Because we originally ignored sleep stage characteristics when selecting the recordings, we had not realized we included 16 recordings with binary wake-sleep annotations. In benchmark v1.1, those recordings have now been replaced with suitable ones where full sleep staging using R&K or AASM has been provided.

## 2. Dataset

### 2.1. Benchmark v1.0

In general, we refer readers to our original paper for full details on the benchmark v1.0 dataset [1]; we reproduce, at times exactly, some of the summary information here for the reader’s convenience.

#### 2.1.1. Goal and source datasets

Our goal was to create a dataset that included as much natural diversity as we could find. Therefore, we used recordings from five large studies in the National Sleep Research Resource [3]:

- The Cleveland Children’s Sleep and Health Study (CCSHS) [6] included 515 pediatric PSGs.
- The Cleveland Family Study (CFS) [7] included 730 PSGs from a wide age range.
- The Childhood Adenotonsillectomy Trial (CHAT) [8] included 1639 pediatric PSGs.
- The Multi-Ethnic Study of Atherosclerosis (MESA) [9] included 2056 PSGs from older subjects.
- The Wisconsin Sleep Cohort (WSC) [10] included 3671 PSGs from middle-aged to older subjects.

These datasets consist of PSG recordings scored using either R&K (CCSHS, CFS, and WSC) or AASM (CHAT and MESA). In addition to the wide range of ages (5 to 90 years), these datasets provide diversity in sex, race, ethnicity, and medical conditions.

#### 2.1.2. ECG data

Even though each recording included data from many biophysical signals, we used only ECG lead I (the limb lead across the heart). Some studies provided ECG lead I as a single channel of ECG data (often labeled “ECG” or “EKG”). Other studies separately provided recordings of the right (RA) and left (LA) limb electrodes. For those studies, we calculated ECG lead I by subtracting the two electrodes from each other (i.e., RA-LA).

#### 2.1.3. Automated pre-processing

Because ECG is often an afterthought for PSG collection, there was a considerable variation in ECG data quality (e.g., intermittent or poor connections, different sampling rates, and environmental noise). To that end, all recordings went through an automated pre-processing algorithm described below. It bears mentioning that we took the recordings as-is and did not trim wake periods before the subject fell asleep (mean sleep latency = 1.3±1 hr.—one SD) or after the subject woke up.

- If the ECG length was not a multiple of 30 seconds, we trimmed it down to the next nearest 30-second epoch length.
- We silenced (i.e., set the signal to a value of 0) sections affected by intermittent connections. If we derived the ECG lead from two electrodes, we silenced both signals whenever there was a connection issue with either.
- If we used two electrodes, we subtracted one from the other to obtain ECG lead I.
- High-pass filter (0.5 Hz) the data to attenuate baseline wander but maintain longer features, such as T waves.
- Remove 60 Hz line noise with a notch filter.
- Remove any additional, automatically-detected, constant-frequency noises using notch filters.
- Resample to a single common frequency (256 Hz).
- Normalize using a robust z-score.

#### 2.1.4. Acceptable recording criteria

We wanted to ensure that all recordings were of decent quality (i.e., an allowance for reasonable quality variations, while excluding “garbage data”). Therefore, we only specified selection criteria based on the ECG data. In other words, none of the original criteria were based on the human-scored sleep stage classification (e.g., time spent in N3, etc.).

1. At least 5 hours of data, but no more than 15 hours.
2. Contain the lead I ECG channel or the two electrode channels necessary to derive it.
3. A sampling rate of at least 100 Hz.
4. There were three or fewer constant-frequency noises (including 60 Hz).
5. At least 85% of the epochs must contain some signal (defined as having more than eight unique values).
6. At least 85% of the epochs must contain at least one template-matching heartbeat.
7. At least 85% of the epochs must contain median absolute deviation (MAD) values ≥ -3 SD of the robust z-scored MAD values for all epochs.
8. The normalization factor must be ≤ 250.
9. After dividing by the normalization factor, ≤ 5% of the data can be extreme values (i.e., values outside ±1).

#### 2.1.5. Recording selection and set building

At this point, there were 5718 recordings remaining (of the original 8611 from all five source studies) that met the above selection criteria. Unfortunately, there were insufficient recordings from subjects in their 3^rd^, 4^th^, 5^th^, and 10^th^ decades to match the U.S. census estimates (decade 1 = age 0- 9yr.). Therefore, we over-sampled subjects from the remaining decades with the following two goals. First, the mean age should match that of the U.S. census. Second, the subjects from the 6^th^ to 9^th^ decades should be over-sampled by the same number. Even with these changes, we still desired to match the sex distributions for each decade, as provided by the census data.

To keep training times reasonable, we selected 4000 recordings that we would use for the training (3000), validation (500), and testing (500) sets. We used random sampling to select the 4000 recordings from the 5718 remaining—with the age and sex distributions specified above. Because there are more recordings than unique subjects, we put additional criteria in the random selection process. First, the 500 recordings in the testing set must come from 500 unique subjects. Furthermore, we allowed more than one recording from the same subject for the training and validation sets on the condition that the subject was only in a single set. Finally, because random selection will probably draw from the original datasets unequally, the last step was to shuffle recordings between the sets to achieve similar ratios of the source datasets across the sets.

### 2.2. Benchmark v1.1

As previously stated, the goal of this update was to remove the 16 recordings that had binary wake-sleep labels (12 from training, 2 from validation, and 2 from testing). To do so, we selected 16 recordings that had age- and sex-matched subjects by requiring the replacement recording to have the same sex, be from the same decade of life, and to meet the other set requirements described above in section 2.1.5. When there was more than one recording that met all of those criteria, the recording that was closest in length was chosen (i.e., similar number of epochs). The updated dataset had the same recording counts as in the original version (3000 for training, 500 for validation, and 500 for testing). Finally, because we drew these 16 from the “remaining set” (that contained 1718 recordings), and had to remove an additional five recordings with wake-sleep labels from that same set, this left 1697 recordings in the “remaining set”.

We aimed to have the randomly selected distribution of subjects match the 2022 U.S. census estimates across decades as well as the mean age (Fig. 1a). Additionally, we mirrored the distribution of age, sex, and recording source across the three sets (i.e., training, validation, and testing). Given the variations in demographics of the source datasets, there are variations in the decade distributions for each study (Fig. 1b). As desired by selecting a broad range of subjects, the chosen recordings contained a wide distribution in stage ratios (i.e., the percent of the night spent in a specific stage), including a substantial number of recordings that had no epochs containing N1, N3, or both (Fig. 1c). Additionally, there is substantial variability in recording lengths, from 5.5 to 14.3 hours (Fig. 1d). When stratifying the recordings by decade, there are noticeable age-dependent shifts in the stage ratios (Fig. 1e). We expected the likelihood of time-dependent changes in stage ratios (e.g., N3 is more common early in the night, and REM more common at the end) and the age-dependent shifts (e.g., N3 decreases with age).

**Fig. 1.**
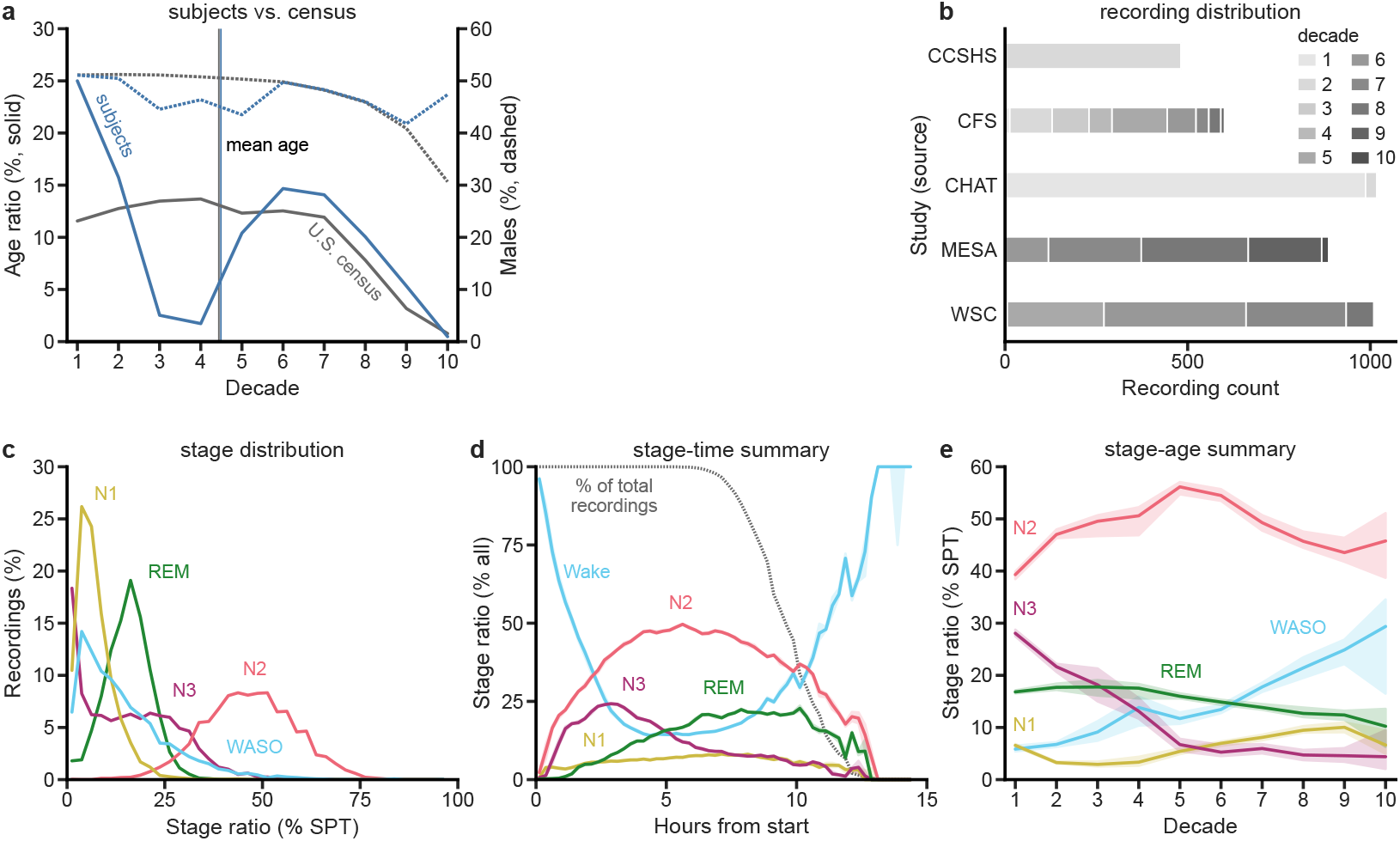
Sleep datasets statistics. This figure reproduces the same panels and captions from the original paper using the data from the updated benchmark dataset v1.1. This figure is nearly indistinguishable from Fig. 1 in the original. (a) We aimed to select subjects (blue lines) to match the U.S. census statistics (gray lines) by age (solid lines) and sex (dashed lines). The lack of subjects in decades 3-5 is a limitation of the available datasets (decade 1=age 0-9yr.), with subjects added to other decades to achieve the same mean age as the census data. (b) The 4000 recordings came from five studies, with the distribution of the subjects’ ages in decades shown and reproduced in Table 1. (c) There is a wide distribution of stage ratios as a percentage of sleep period time (SPT, i.e., the period between and including the first and last epoch of sleep). Wake after sleep onset (WASO) is any wake (arousals) during SPT. (d) As expected from previous studies, recordings show time-dependent changes in the relative proportion of the various sleep stages. E.g., N3 is more common at the beginning, and REM is more common at the end. (e) The data also show expected age-dependent changes in sleep. In particular, the ratio of time spent in stage N3 declines with age, whereas arousals (WASO) increase.

Table 1 details which source dataset the removed and added recordings were from. Next, Table 2 outlines the epoch counts for each set in the dataset. Finally, Table 3 shows a comparison of the two benchmarks of the recording counts that contain specific sleep stage combinations.

**Table 1.** Recording source dataset counts.

| Dataset | Total | Source dataset |  |  |  |  |
| --- | --- | --- | --- | --- | --- | --- |
|  |  | CCSHS | CFS | CHAT | MESA | WSC |
| Benchmark v1.0 | 4000 | 482 | 594 | 1019 | 896 | 1009 |
| Removed | 16 | 0 | 3 | 2 | 11 | 0 |
| Added | 16 | 0 | 10 | 2 | 1 | 3 |
| Benchmark v1.1 | 4000 | 482 | 601 | 1019 | 886 | 1012 |

**Table 2.** Epoch counts.

| Set | Wake | Epoch counts for each stage |  |  |  |  | Total |
| --- | --- | --- | --- | --- | --- | --- | --- |
|  |  | N1 | N2 | N3 | REM | Unscored |  |
| Training | 979,199 | 210,205 | 1,313,052 | 457,909 | 449,020 | 41,462 | 3,450,847 |
| Validation | 165,580 | 34,998 | 217,695 | 74,150 | 72,150 | 7,223 | 571,796 |
| Testing | 165,179 | 34,693 | 219,713 | 74,401 | 73,382 | 7,209 | 574,577 |
| Total | 1,309,958 | 279,896 | 1,750,460 | 606,460 | 594,552 | 55,894 | 4,597,220 |

**Table 3.**
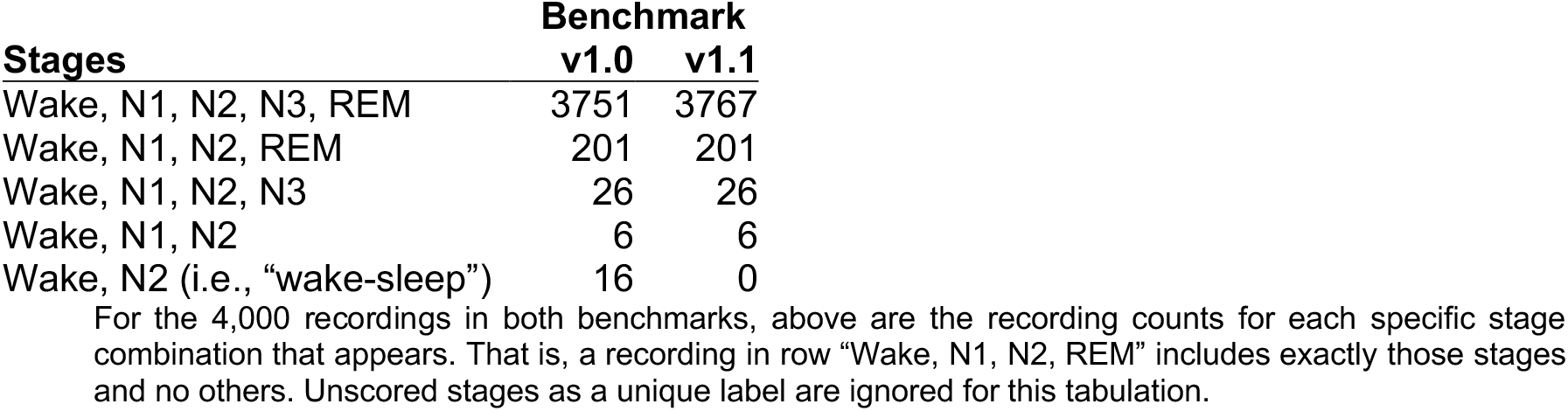
Sleep stage combination recording counts.

## 3. Conclusion

Benchmark v1.1 updates the previously released sleep stage classification dataset by replacing 16 recordings that contained only binary wake-sleep annotations with age- and sex-matched recordings scored using either the R&K or AASM standards. The total number of recordings and the composition of the training, validation, and testing sets remain unchanged. All other dataset selection criteria and distributions are preserved. This update corrects an ontological oversight present in benchmark v1.0 and provides a revised dataset for use in full sleep stage classification tasks.

## Data Availability

All human input data for this study came from the datasets listed and are available at the National Sleep Research Resource (https://sleepdata.org/). We have provided set filenames on GitHub at (https://github.com/adammj/ecg-sleep-staging/).

https://github.com/adammj/ecg-sleep-staging/

## Data availability

All human input data for this study came from the datasets listed and are available at the National Sleep Research Resource (https://sleepdata.org/).

## Code availability

We have provided set filenames on GitHub at (https://github.com/adammj/ecg-sleep-staging/).

## Author contributions

A.M.J. updated the dataset and wrote the manuscript. B.R.S. provided feedback on the text. Both authors contributed to the final manuscript.

## Acknowledgments

The Cleveland Children’s Sleep and Health Study (CCSHS) was supported by grants from the National Institutes of Health (RO1HL60957, K23 HL04426, RO1 NR02707, M01 Rrmpd0380-39). The National Sleep Research Resource was supported by the National Heart, Lung, and Blood Institute (R24 HL114473, 75N92019R002).

The Cleveland Family Study (CFS) was supported by grants from the National Institutes of Health (HL46380, M01 RR00080-39, T32-HL07567, RO1-46380). The National Sleep Research Resource was supported by the National Heart, Lung, and Blood Institute (R24 HL114473, 75N92019R002).

The Childhood Adenotonsillectomy Trial (CHAT) was supported by the National Institutes of Health (HL083075, HL083129, UL1-RR-024134, UL1 RR024989). The National Sleep Research Resource was supported by the National Heart, Lung, and Blood Institute (R24 HL114473, 75N92019R002).

The Multi-Ethnic Study of Atherosclerosis (MESA) Sleep Ancillary study was funded by NIH-NHLBI Association of Sleep Disorders with Cardiovascular Health Across Ethnic Groups (RO1 HL098433). MESA is supported by NHLBI funded contracts HHSN268201500003I, N01-HC-95159, N01-HC-95160, N01-HC-95161, N01-HC-95162, N01-HC-95163, N01-HC-95164, N01-HC-95165, N01-HC-95166, N01-HC-95167, N01-HC-95168 and N01-HC-95169 from the National Heart, Lung, and Blood Institute, and by cooperative agreements UL1-TR-000040, UL1-TR-001079, and UL1-TR-001420 funded by NCATS. The National Sleep Research Resource was supported by the National Heart, Lung, and Blood Institute (R24 HL114473, 75N92019R002).

This Wisconsin Sleep Cohort Study (WSC) was supported by the U.S. National Institutes of Health, National Heart, Lung, and Blood Institute (R01HL62252), National Institute on Aging (R01AG036838, R01AG058680), and the National Center for Research Resources (1UL1RR025011). The National Sleep Research Resource was supported by the U.S. National Institutes of Health, National Heart Lung and Blood Institute (R24 HL114473, 75N92019R002).

